# Comparative analysis of pediatric Respiratory Syncytial Virus epidemiology and clinical severity before and during the COVID-19 pandemic in British Columbia, Canada

**DOI:** 10.1101/2022.11.18.22282477

**Authors:** Marina Viñeta Paramo, Bahaa Abu-Raya, Frederic Reicherz, Rui Yang Xu, Jeffrey N. Bone, Jocelyn A. Srigley, Alfonso Solimano, David M. Goldfarb, Danuta M. Skowronski, Pascal M. Lavoie

## Abstract

**Background:** The COVID-19 pandemic affected Respiratory Syncytial Virus (RSV) circulation and surveillance, causing logistical complexity for health systems. Our objective was to describe changes in epidemiology and clinical severity of RSV cases in British Columbia, Canada.

**Methods:** Comparative analysis of RSV detections in children <36 months at BC Children’s Hospital (BCCH) between September 1 and August 31 of 2017-18, 2018-19, 2019-20, 2020-21 and 2021-22.

**Results:** About one-fifth of children tested RSV positive on average across all periods. The median age of RSV cases was 11.8 [IQR: 3.8–22.3] months in 2021-22 versus 6.3 [IQR: 1.9–16.7] months in 2017-20 (p<0.001). Increased testing in 2021-22 (n=3,120) compared to 2017-20 (average n=1,222/period) detected milder infections with lower proportion hospitalized in all age subgroups <6 (26.0%), 6-11 (12.3%), 12-23 (12.2%) and 24-35 (16.0%) months versus 2017-20 (49.3%, 53.5%, 62.6%, 57.5%, respectively) (all p<0.001). Children <6 months consistently comprised most hospitalizations and those born prematurely <29 weeks or with chronic respiratory co-morbidities remained at highest hospitalization risk in 2021-22. Among hospitalized cases, intensive care, respiratory support or supplemental oxygen use did not differ between the 2017-20 and 2021-22 periods.

**Conclusions:** RSV circulation halted during the pandemic, but with the lifting of mitigation measures a subsequent resurgence in children <36 months of age was accompanied by shift toward older (24-35 month) cases in 2021-22, without increased severity. For the 2022-23 period, increased circulation and residual vulnerability in additional birth cohorts spared from RSV infection during the pandemic could have marked cumulative healthcare impact, even without increase in proportion hospitalized.

## Introduction

Respiratory Syncytial Virus (RSV) is a leading cause of morbidity and mortality in young children globally (1). Before the coronavirus disease 2019 (COVID-19) pandemic, RSV cases peaked predictably every winter between September and April in Canada (2). During the pandemic, RSV infections dramatically declined internationally, losing the typical seasonal pattern in many regions of the world (3–5). In Canada, 986 RSV cases were reported between September 2020 and August 2021, compared to the average 18,525 cases between September and August of the prior 2017-18, 2018-19 and 2019-20 periods (6). These drastic changes in RSV epidemiology, which were also observed for most respiratory viruses (2), may be attributed to COVID-19 mitigation strategies, or, according to others, viral interference from SARS-CoV-2 (7). Subsequently, the relaxing of population-level pandemic mitigation strategies coincided with a resurgence of RSV cases with atypical seasonality (8–11). The eastern Canada province of Quebec saw a most atypical resurgence of cases during the summer. Anecdotally, the BC Children’s Hospital (BCCH) Pediatric Emergency Department experienced a rise in RSV cases in the fall/winter of 2021-22 (12). However, the severity of these cases remains unclear as, to the best of our knowledge, no previous study reported hospitalizations. A better understanding of the epidemiology and clinical features of the resurgence is crucial to help anticipate subsequent RSV seasons, and inform RSV prevention programs health resource planning.

Newborns are immunologically naïve and depend on maternal antibodies transferred during pregnancy to provide protection against respiratory viruses. After birth, transplacental passively-transferred maternal IgG antibodies wane gradually within 4 to 6 months. By two years of age, most children have seroconverted to RSV through infection (13,14). A previous study estimated that RSV immunity in the population could wane over 6 to 12 months, based on biennial changes in RSV cases in Western Canada (15). Moreover, we reported a waning of serum RSV-neutralizing antibodies in women and young children in the Vancouver metropolitan area between February-June 2020 and May-June 2021, in the context of reduced RSV exposure (16). A prolonged lack of RSV in children born immediately before and during the pandemic may result in residual susceptibility among spared cohorts upon RSV resurgence, with uncertain implications for severity and strain upon healthcare system capacity.

The objective of this study was to describe the epidemiology and compare the clinical severity of RSV cases in children younger than 36 months at BCCH before and during the COVID-19 pandemic.

## Methods

### Study design

This population-based analysis summarizes laboratory-confirmed RSV cases in BC, comparing the epidemiology and severity of detections among children younger than 36 months diagnosed at BCCH during five one-year periods between September 1^st^ to August 31^st^ of 2017-18, 2018-19, 2019-20, 2020-21 and 2021-22.

### Setting

BCCH is located in Vancouver, BC, and is the main pediatric referral center for children up to 17 years of age for the province (∼5.2 million provincial population in 2022, including ∼180 thousand children under 36 months of age and ∼871 thousand children up to 17 years of age (17)). The BCCH pediatric emergency department sees >49,000 patients annually and its Pediatric Intensive Care Unit (PICU) is the largest in the province with 22 beds, providing >80% of all provincial pediatric intensive care needs (18). The BCCH Microbiology Laboratory performs respiratory virus testing using two diagnostic nucleic acid amplification test panels, both including RSV (**Supplemental table 1**). Testing criteria include symptomatic children with risk factors for a severe respiratory infection, nosocomial infection, or travel outside of Canada. In April 2020, testing guidelines were updated to include SARS-CoV-2 and broader respiratory viral testing indications specifically for children admitted to hospital.

### population

Children <36 months who tested positive for RSV at BCCH between September 1^st^, 2017, and August 31^st^, 2022 were identified from weekly reports of all respiratory specimens performed at the BCCH Microbiology Laboratory. Tests for the same subject with the same result within 4 weeks were merged, with the earliest record retained as date of diagnosis. Subjects were excluded from severity analysis if their electronic medical records could not be retrieved. To estimate potential biases, RSV data for all of BC were obtained from the Canadian Respiratory Virus Detection Surveillance System reports (2), which includes >99% of RSV cases diagnosed at BCCH.

### Data collection

The following data were systematically collected from BCCH electronic medical charts: sex, age, gestational age, medical comorbidities, first three digits of residency postal code, date and result of respiratory viral testing (date, viruses detected, including RSV, and panel used), and data from RSV-related hospitalizations for at least 24h (with length of stay, use, type and length of respiratory therapy, admission to ICU and RSV-related in-hospital mortality).

### Statistical analysis

RSV positivity rates were calculated as number of children testing positive for RSV divided by number of children tested. Unless otherwise specified, the number of children tested was used as the denominator for all other analyses. To compare RSV cases throughout the study period, we defined RSV one-year periods occurring between September 1^st^ (the month typically immediately preceding the beginning of the RSV peak season (19)) and August 31^st^ to provide a more complete, uninterrupted picture up to the following year. Baseline and clinical data from 2021-22 were compared to data from the corresponding September 1^st^ to August 31^st^ 2017-18, 2018-19 and 2019-20 periods using a Mann Whitney U test for numerical data, and binomial logistic regression with Wald’s test, including odds ratio (OR) and confidence intervals (CI), for categorical data. P-values <0.05 were considered statistically significant. Subgroup analyses were also performed to study clinical severity differences by age groups (i.e., <6, 6 to 11, 12 to 23, and 24 to <36 months old). For simplicity, the latter three periods are referred to as pre-pandemic, also because we expected negligible RSV cases towards the end of March 2020 after the COVID-19 pandemic was declared, on March 11, 2020, by the WHO, and strict lockdown mitigation measures were initiated in BC by the middle of March 2020.

To investigate a possible change in risk factors for RSV-related hospitalization and ICU admission in 2021-22, the effects of sex, postnatal age (i.e., <12 months), prematurity (i.e. born <29 weeks of gestation) and types of respiratory comorbidities (based on the American Academy of Pediatrics criteria for palivizumab administration (20)) were included in the analysis. Binary logistic regression models were performed for each variable comparing the 2021-22 period to the pooled 2017-20 periods. All risk factors were included in a multivariable logistic regression analysis, excluding missing data. OR and CI were calculated as exponentiated coefficients from the logistic regression models; p-values were obtained with Wald’s test. Statistics were calculated using R v4.1.3 and GraphPad Prism v9.3.1.

### Ethics

This study was approved by the University of British Columbia Children’s & Women’s Research Ethics Board, who also waived the requirement for participants’ consent since no identifiable data was required or used during this study (approval number H11-03424).

## Results

### RSV cases at BCCH

A total of 11,069 RSV tests were performed at BCCH between September 1^st^, 2017 and August 31^st^, 2022, of which 8,687 tests were performed in 7,867 children younger than 36 months. Of these, 1,556 RSV tests were positive in 1,496 children (**Supplemental Figure 1**).

Children <36 months with RSV at BCCH comprised 810/4,894 (16.6%) of all cases diagnosed in BC in 2017-20, 1/8 (12.5%) in 2020-21 and 685/8,194 (8.4%) in 2021-22. RSV cases at BCCH showed similar seasonality to BC overall (**Figure 1**), peaking in December or January. Among BCCH detections, the peak in absolute cases was earlier by 7 weeks and in test-positivity by 3 weeks in 2021-22 compared to the average of 2017-20.

**Figure 1:**
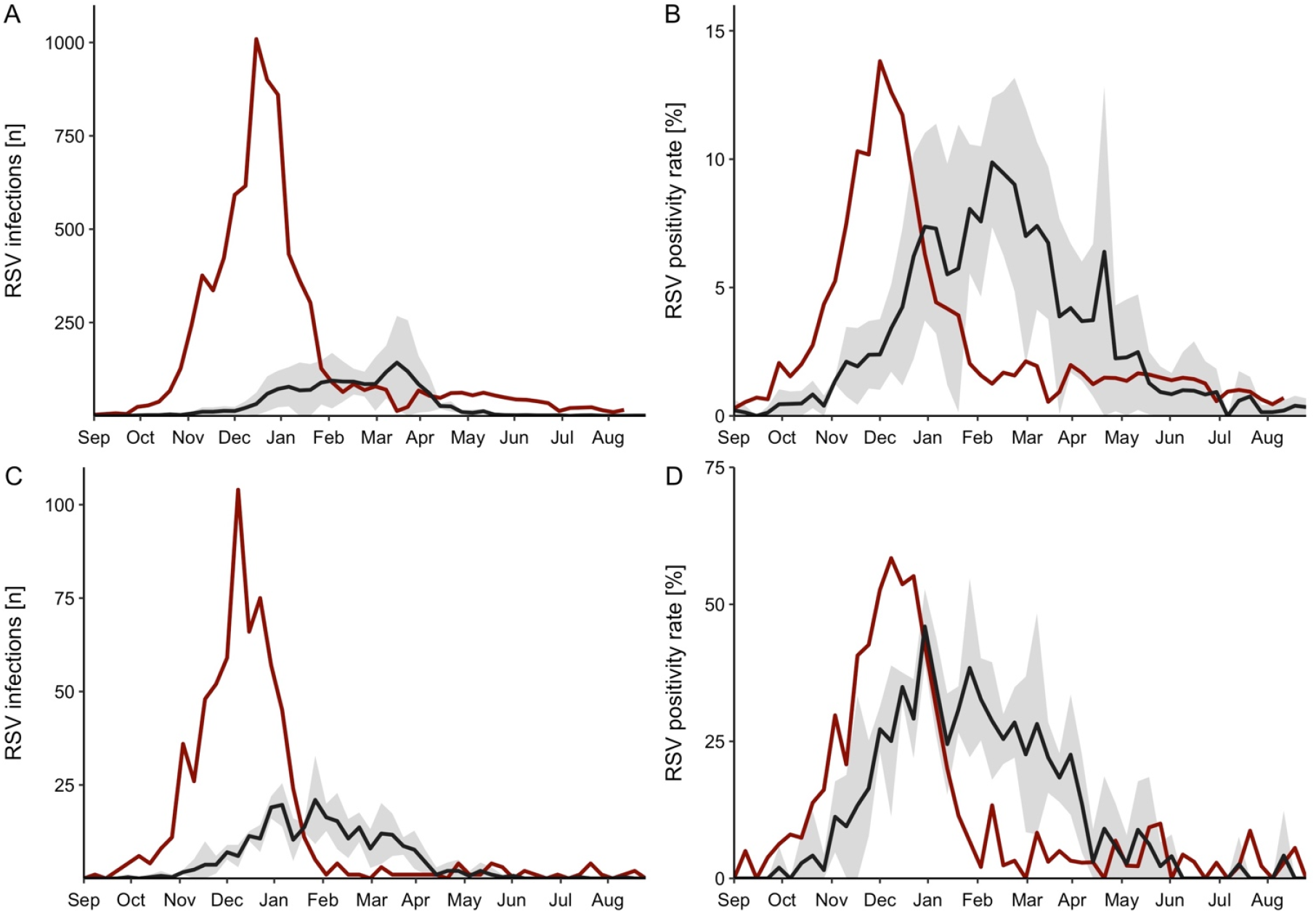
Overall RSV cases reported in BC and in children younger than 36 months at the BC Children’s Hospital (BCCH). Weekly data referred to the date of respiratory specimens collected, including laboratory-confirmed RSV cases and testing positivity rates in BC (A, B) and at BCCH (C, D). Data from BC (A, B) were compiled from the *Canadian Respiratory Virus Detection Surveillance System* weekly reports publicly available from the Public Health Agency of Canada^2^. The black line represents averaged data from September 1^st^ to August 31^st^ in 2017-18, 2018-19 and 2019-20, with 95% confidence intervals (gray area). Red line represents data between September 1^st^, 2021 and August 31^st^, 2022. Data from September 1^st^, 2020 and August 31^st^, 2021 are excluded from the figure, as there were too few RSV cases during this period (8 in BC and 1 at BCCH). Monthly intervals (x-axis) based on week for the first day of the month from the 2021-22 calendar. Plots A&B are missing data on week 51 in 2021, therefore the line is connecting values between week 50 and week 52.

Of an average of 1,222 children <36 months who were tested for RSV at BCCH for each one-year period in 2017-20, an average of 270 (22.1%) were positive, including 306/1,206 (25.4%) in 2017-18, 283/1,281 (22.1%) in 2018-19 and 221/1,179 (18.7%) in 2019-20. RSV detection was much lower in 2020-21 at just 1/1,081 (0.1%), increasing again to 685/3,120 (22.0%) during the subsequent 2021-22 period (**Supplemental table 2**). The test-positivity was significantly greater than the 2020-21 (p<0.001) and the 2019-20 (p=0.015) periods, did not significantly differ from the 2018-19 period (p=0.921) and resulted significantly lower than the 2017-18 period (p=0.016).

The median age of children <36 months with RSV at C&W in 2021-22 was higher (11.8 months [IQR: 3.8 - 22.3]) compared to 2017-20 (6.3 [IQR: 1.9 – 16.7]) (p<0.001) (**Table 1**; **Supplemental figure 2**). Children 24 to <36 months represented a larger proportion of RSV cases in 2021-22 (21.9%) compared to the combined 2017-20 periods (11.5%) (p<0.001) (**Table 1**). RSV test-positivity rates were comparable between periods for the <6, 6-11 and 12-23 months. However, in 2021-22, the age sub-group 24-<36 months showed statistically significantly higher RSV test-positivity rates (150/599; 25.0%) compared to 2018-19 (31/190; 16.3%), 2019-20 (26/178; 14.6%), and 2020-21 (1/151; 0.7%) (all p<0.05). Although also higher in 2021-22 than in 2017-18 (35/170; 20.6%), without reaching statistical significance (p=0.211) (**Table 2**).

**Table 1:** Baseline characteristics of children younger than 36 months with RSV at BCCH, by one-year period.

| **Characteristic** | **September 1<sup>st</sup> to August 31<sup>st</sup> period** (total # RSV cases; n = 1,495) |  |  |  |  | **P value<sup>2</sup>** |
| --- | --- | --- | --- | --- | --- | --- |
|  | **2017-18**<br>(N = 306) | **2018-19**<br>(N = 283) | **2019-20**<br>(N = 221) | **2017-20**<br>**combined**<br>(N = 270)<sup>1</sup> | **2021-22**<br>(n = 685) |  |
| Age at diagnosis (months), median (IQR) | 5.3<br>(1.8 - 15.9) | 6.9<br>(1.9 - 17.3) | 7.1<br>(2.5 - 16.2) | 6.3<br>(1.9 - 16.7) | 11.8<br>(3.8 - 22.3) | <0.001 |
| Resident from Vancouver metro area<sup>3</sup>, n (%) | 257 (84.0) | 255 (90.1) | 194 (87.8) | 235 (87.0) | 648 (94.6) | <0.001 |
| Age < 6 months, n (%) | 161 (52.6) | 136 (48.1) | 104 (47.1) | 134 (49.5) | 241 (35.2) | <0.001 |
| 6 to 11 months, n (%) | 45 (14.7) | 41 (14.5) | 33 (14.9) | 40 (14.7) | 106 (15.5) | 0.992 |
| 12 to 23 months, n (%) | 65 (21.2) | 74 (26.1) | 58 (26.2) | 66 (24.3) | 188 (27.4) | 0.121 |
| 24 to 35 months, n (%) | 35 (11.4) | 32 (11.3) | 26 (11.8) | 31 (11.5) | 150 (21.9) | <0.001 |
| Sex, n female (%) | 142 (46.4) | 122 (43.1) | 93 (42.1) | 119 (44.1) | 305 (44.5) | 0.688 |

<sup>1</sup> N (%) corresponds to the average, combining data from 3 one-year period (2017-20), rounded to the nearest unit;
<sup>2</sup> Comparing the data from the combined 2017-20 periods versus 2021-22, using a Wald test on binomial logistic regression for proportions (i.e., age category, sex and residency area) and a Mann-Whitney U test for age;
<sup>3</sup> Classified as the regional district of metro Vancouver, including 23 local authorities (linked with 3 first digits of postal code).

**Table 2:**
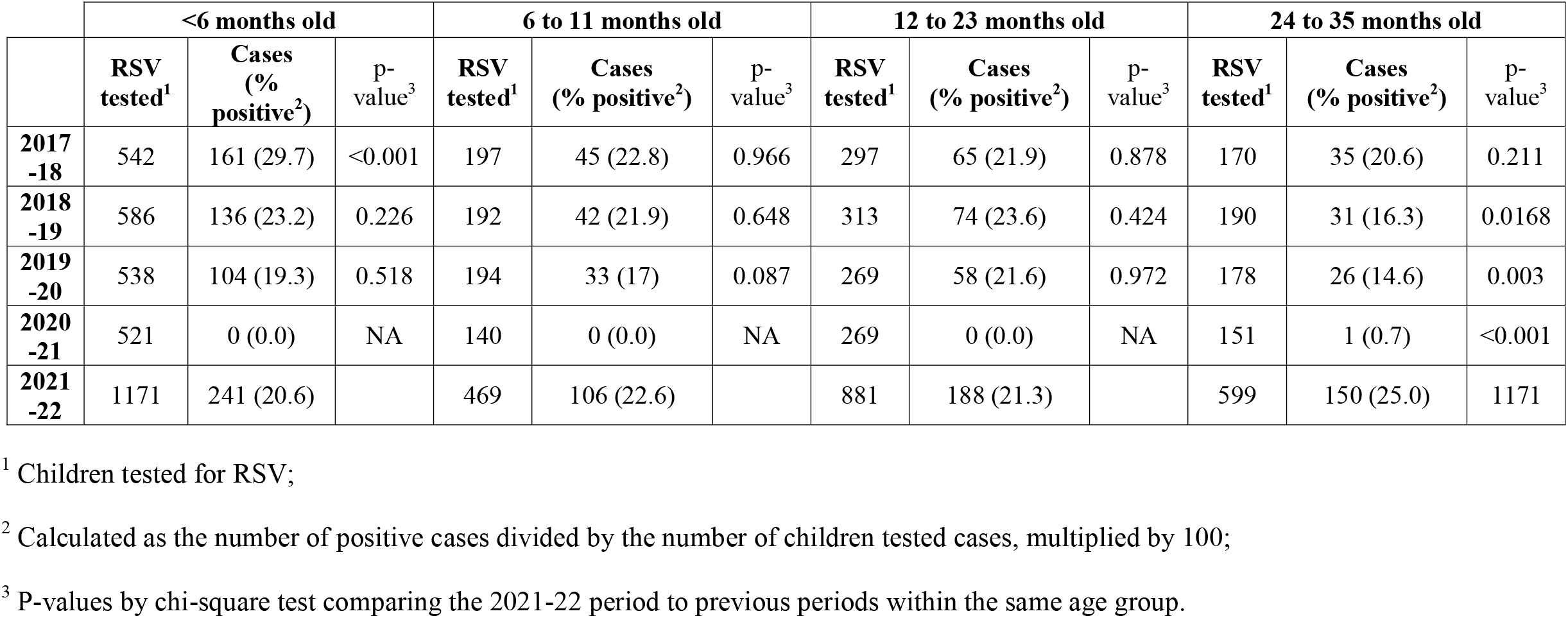
Analysis of RSV cases and positivity rates by age groups in children younger than 36 months at BCCH, by one-year period.

### Clinical severity analysis

Of 1,496 children <36 months of age with RSV at BCCH, 59 lacked sufficient data for analysis of clinical severity. Excluding the one case in 2020-21, this yielded 1,436 (96.0%) children for analysis (**Supplemental figure 1**).

There were no significant differences in the proportion of RSV cases from children with comorbidity or born prematurely between periods (**Table 3**; p>0.050).

**Table 3:**
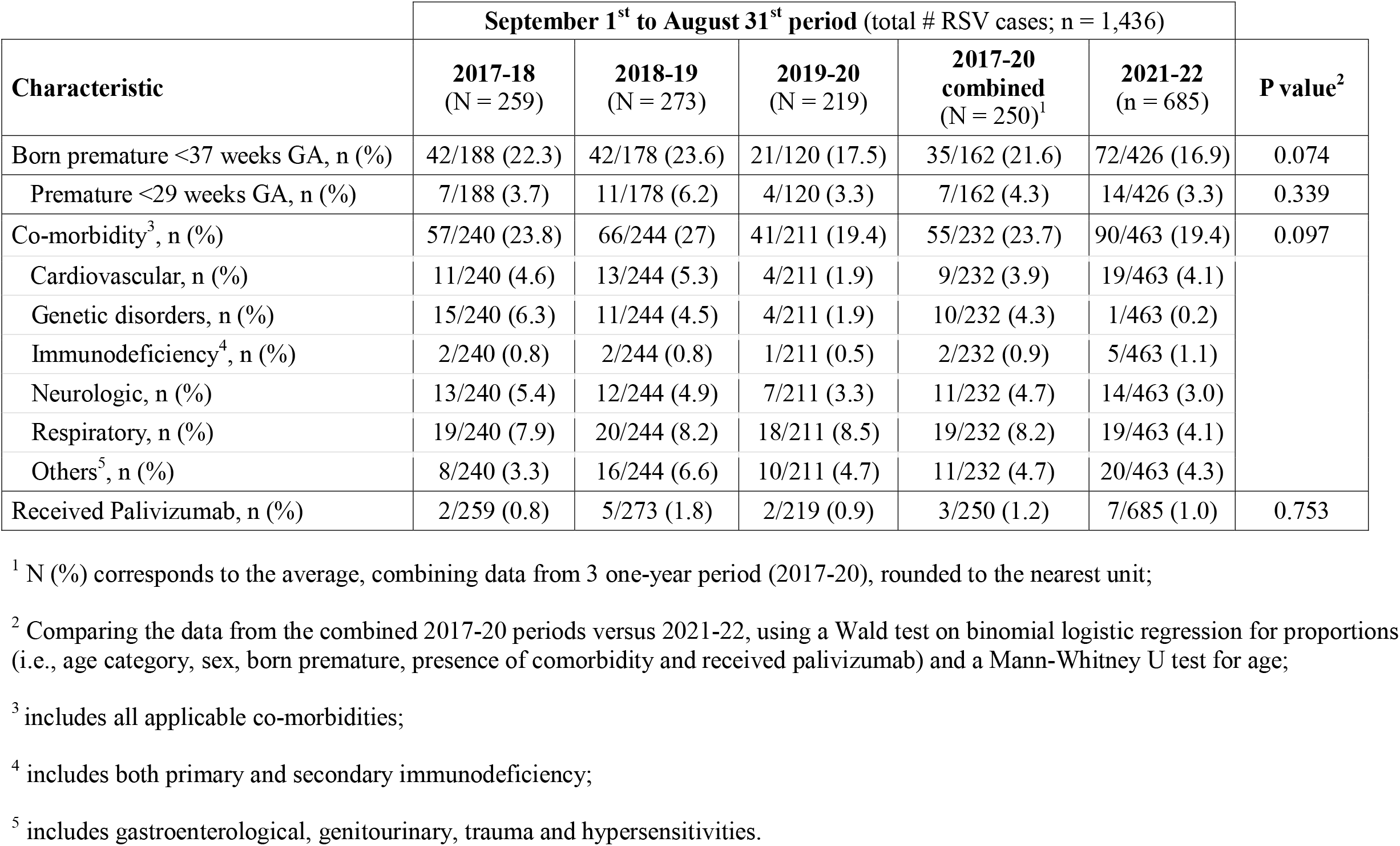
Co-morbidities in children younger than 36 months with RSV at BCCH, by one-year period.

The percentage of children <36 months hospitalized for RSV at BCCH was lower in 2021-22 (123/685; 18.0%) compared to the averaged 2017-20 periods (135/250; 54.1%; p<0.001) or to any of the periods separately: 153/259; 59.1% in 2017-18, 138/273; 50.5% in 2018-19 and 115/219; 52.5% in 2019-20 (all p<0.001; not shown) (**Table 4**). Similarly, in the age sub-group analysis, the percentage of RSV-related hospitalizations in 2021-22 was significantly lower in all age groups compared to all preceding periods or the averaged 2017-20 (all p<0.001) (**Supplemental table 3**).

**Table 4:** Severity outcomes for children under 36 months of age with RSV at BCCH, by one-year period.

|  | **September 1<sup>st</sup> to August 31<sup>st</sup> period** (total # RSV cases; n = 1,436) |  |  |  |  |  |  |
| --- | --- | --- | --- | --- | --- | --- | --- |
| **Severity outcome** | **2017-18**<br>(n = 259) | **2018-19**<br>(n = 273) | **2019-20**<br>(n = 219) | **2017-20 periods**<br>**combined** (n = 250)<sup>1</sup> | **2021-22**<br>(n = 685) | **Effect estimates**<br>**(OR, CI)<sup>2</sup>** | **P value<sup>2</sup>** |
| Hospitalized, n (%) | 153 (59.1) | 138 (50.5) | 115 (52.5) | 135 (54.1) | 123 (18.0) | 0.18 (0.14 – 0.23) | <0.001 |
| Length of in-hospital stay (days),<br>median (IQR) | 4 (2 - 6) | 4 (2-5) | 3 (2-5) | 3 (2 - 6) | 3 (2 - 5) | Not applicable | 0.077 |
| Intensive Care Unit (ICU)<br>admission,<br>n (% hospitalized) | 37 (24.2) | 25 (18.1) | 22 (19.1) | 28 (20.7) | 27 (22.0) | 1.02 (0.62 – 1.67) | 0.970 |
| Supplemental O<sub>2</sub><sup>3</sup>, n (%<br>hospitalized) | 102 (66.7) | 93 (67.4) | 75 (65.2) | 90 (66.7) | 91 (74.0) | 1.46 (0.94 – 2.33) | 0.143 |
| Mechanical ventilation, n (%<br>hospitalized) | 20 (13.1) | 7 (5.1) | 9 (7.8) | 12 (8.9) | 14 (11.4) | 0.69 (0.25 – 1.60) | 0.381 |
| RSV-related death, n (%<br>hospitalized) | 0 (0) | 2 (1.4) | 0 (0) | 1 (0.5) | 1 (0.8) | Not tested | - |

<sup>1</sup> N (%) corresponds to average number per annual period, combining data from the 2017-20 period except for length of in-hospital stay where median and IQR were calculated from all values from the 2017-20 period;
<sup>2</sup> Comparing the combined data from the 2017-20 periods versus the 2021-22 period, using a Wald test on binomial logistic regression for proportions (i.e., hospitalization, supplemental oxygen, ICU admission, mechanical ventilation, and mortality) with odds ratio (OR) and 95% confidence interval (CI) reported and Mann-Whitney U test for length of in-hospital stay;
<sup>3</sup> Supplemental oxygen included patients receiving blow-by oxygen, nasal prongs with low flow O<sub>2</sub>, high flow nasal prongs, bilevel positive airway pressure (BiPAP) and continuous positive airway pressure (CPAP). Frequencies for each method reported in **Supplemental table 3**.

Children <6 months comprised about half of all RSV hospitalizations annually at BCCH, with no difference between 2021-22 (63/123; 51.2%) and the 2017-20 periods (61/135; 45.1%) (p=0.214).

Among all children hospitalized at BCCH, the proportion requiring ICU admission, supplemental oxygen, or mechanical ventilation did not differ significantly in 2021-22 compared to the averaged 2017-20 periods (22% vs. 21%; 74% vs. 67%, 11% vs. 9%, respectively) (all p > 0.050) (**Table 4**; **Supplemental table 3** and **Supplemental table 4**). Mortality was rare across the study, with two RSV-related in-hospital deaths during the 2018-19 period (one <6 months and another 12-23 months old), and a third during the 2021-22 period (12-23 months old) (**Table 4**).

### Risk factor analysis

In multivariable analyses to assess independent factors contributing to the likelihood of hospitalization among RSV cases during the 2017-20 periods, male sex (OR: 0.64; 95%CI 0.41 – 0.98; p=0.044) and age <12 months (OR 0.33; 95%CI: 0.18 – 0.58; p=0.001) were less often associated with hospitalization. This possibly reflects greater testing proclivity inclusive of milder infections among the very young targeted for testing per routine guidance because considered at higher risk (**Table 5**). In the 2021-22 period age and sex were not associated with RSV-related hospitalization, but premature birth below 29 weeks of gestation (OR: 4.07; 95%CI 1.26 – 14.34; p=0.020) and presence of chronic respiratory conditions (OR: 4.75; 95%CI: 1.54 – 16.25; p=0.008) were associated with greater likelihood of being hospitalized (**Table 5**). Multivariable analysis of grouped comorbidities indicated that respiratory morbidity was the most and only comorbidity significantly associated with RSV hospitalizations (OR: 8.05; 95%CI: 2.84 - 24.54) (**Supplemental table 5**).

**Table 5:** Risk factor analysis for an RSV-related hospitalization and ICU admission in children under 36 months of age at BCCH.

|  | Univariate analysis |  | Multivariate analysis |  |
| --- | --- | --- | --- | --- |
|  | 2017-20 periods combined<br>(n = 732) | 2021-22<br>(n = 676) | 2017-20 periods combined (n = 732) | 2021-22<br>(n = 676) |
|  | OR (95%CI) | OR (95%CI) | OR (95%CI) | OR (95%CI) |
| **Hospitalization<sup>1</sup>** |  |  |  |  |
| Sex<sup>3</sup> | 0.86 (0.60 – 1.22) | 1.09 (0.74 - 1.62) | **0.64 (0.41 – 0.98)** | 0.93 (0.60 – 1.46) |
| Younger than 12 months old | 0.62 (0.45 – 0.84) | 1.73 (1.17-2.60) | **0.33 (0.18 - 0.58)** | 1.22 (0.77 – 1.96) |
| Prematurity <29 weeks GA | 7.20 (1.48 - 129.94) | 5.0 (1.70 - 16.73) | 2.41 (0.44 - 44.90) | **4.07 (1.26 – 14.33)** |
| Respiratory comorbidities<sup>4</sup> | 5.22 (2.47 - 12.81) | 4.95 (1.80 - 14.86) | 5.05 (0.98 – 92.78) | **4.75 (1.54 – 16.25)** |
| **ICU admission<sup>2</sup>** |  |  |  |  |
| Sex<sup>3</sup> | 0.82 (0.51 - 1.33) | 1.27 (0.53 - 3.17) | 0.85 (0.49 – 1.45) | 1.20 (0.45 – 3.33) |
| Younger than 12 months old | 0.97 (0.60 - 1.59) | 1.90 (0.75 – 5.26) | 1.16 (0.64 – 2.17) | **4.08 (1.28 – 17.02)** |
| Prematurity <29 weeks GA | 3.07 (1.16 – 7.74) | 5.72 (1.39 – 25.09) | **3.40 (1.18 – 9.62)** | **6.69 (1.28 – 39.41)** |
| Respiratory comorbidities<sup>4</sup> | 1.07 (0.49 - 2.19) | 2.76 (0.66 – 10.51) | 0.90 (0.30 – 2.37) | 3.02 (0.47 – 18.00) |

<sup>1</sup> Reference group for this analysis is RSV cases not requiring hospitalization;
<sup>2</sup> Reference group for this analysis is RSV cases requiring hospitalization without admission to ICU;
<sup>3</sup> Study group as male and reference group as female;
<sup>4</sup> Respiratory comorbidities were classified from electronic medical charts reporting chronic lung disease, bronchopulmonary dysplasia, congenital airways malformations, cystic fibrosis and possible asthma;
Results with statistical significance in the multivariate analysis are shown in **bold**.

Among hospitalized cases, likelihood of ICU admission was greater for premature infants below 29 weeks of gestation in both 2021-22 (OR: 6.69; 95%CI: 1.28 – 39.41; p=0.026) and 2017-20 periods (OR: 3.40; 95%CI: 1.18 – 9.62; p=0.019). Hospitalized children younger than one year were more likely to be admitted to the ICU during the 2021-22 period (OR: 4.08; 95%CI: 1.28-17.02; p=0.029) but age was not contributing towards ICU admission in 2017-20 (OR: 1.16; 95%CI: 0.64-2.17; p=0.625) (**Table 5**).

## Discussion

Through detailed epidemiological and clinical examination, our study provides greater insight into surveillance observations for RSV in the context of the COVID-19 pandemic, associated mitigation measures and respiratory virus testing and monitoring. In particular, RSV virtually disappeared in BC from September 1, 2020 through August 31, 2021, consistent with observations elsewhere indicating that pandemic control measures had reduced the circulation of respiratory viruses (6). Conversely, enhanced RSV testing during the same period for 2021-22 identified RSV resurgence, including milder cases and more children within the 24-35 month age group, likely spared earlier infection in infancy. Our analysis indicates that this resurgence was not associated with worse outcomes in children <36 months thus far, but that the usual risk factors for RSV severity still applied: namely prematurity, age <12 months, and underlying respiratory comorbidity. In combination, these findings have ongoing implications for RSV preparedness in upcoming seasons, testing and for preventive measures such as palivizumab as well as vaccines, and indications for immunoprophylaxis interventions in high-risk groups, including the use of long-acting monoclonal antibodies for which approval is imminent.

Our findings support anecdotal reports from the BCCH Pediatric Emergency Department indicating a rise in RSV cases in 2021-22 (12). In the wake of pandemic messaging, parents with heightened awareness of even mild acute respiratory illness and with limited primary care options otherwise during the pandemic, may have been more likely to attend emergency rooms, with enhanced testing more likely to find RSV cases than previously. Despite increased testing, in 2021-22 RSV test-positivity rates remained stable compared to prior periods, except for a significant increase among children 24-35 months as also reported by others (8,9,21,22). Both higher absolute detections and greater test-positivity among those 24-35 months suggest a true increase in residual vulnerability in these toddlers in 2021-22 compared to prior periods. In particular, the lack of exposure to RSV during the pandemic might have induced a temporary cohort effect among children spared the usual infection earlier in infancy, prolonging an immunologically naïve state to RSV and delaying their age at first RSV infection. For the upcoming 2022-23 season, increased circulation and residual vulnerability in additional birth cohorts also spared exposure and infection during the pandemic, could tax an already strained healthcare system even with the same proportion by age hospitalized as in other seasons.

Modelling studies had predicted a resurgence of severe RSV cases (23–25). In contrast, our surveillance data showed increased RSV cases without increased severity. These findings are consistent with another study from France, similarly reporting no changes in RSV severity in 2021 (21). A recent study from England that included hospitalizations and emergency department visits confirmed an early pre-season rise in cases and ED visits post-relaxing of pandemic measures. Interestingly, at the same time this study reported a relatively small (10.7% over expected) increase in RSV-related hospital admissions in 2021-22 (26). Since other jurisdictions likely also applied multiplex testing inclusive of SARS-CoV-2 and other respiratory targets (notably RSV and influenza viruses) in an expanded way during the pandemic, the generalizability of our observations warrants assessment elsewhere. In differing from model predictions, our findings reinforce the importance of validating simulations with empirical data including clinical severity outcomes and hospitalization data to better inform pandemic-related forecasting, and measures to avert extraordinary impacts on healthcare resources. For example, to lessen pressure on laboratories and the acute care setting during the upcoming respiratory virus season, targeted testing might be more strategically resumed and/or redirected to less critical care settings and with parents also provided explicit guidance on when they should seek emergency care.

The finding that risk factors for severe infections remained comparable between periods is important. Post-relaxation of physical distancing measures, children <6 months and those born prematurely or with chronic respiratory conditions still represented the bulk of hospitalized children and remained at highest risk for severe disease. The potential role of maternal antibodies in preventing symptomatic or severe RSV infections in very young children warrants consideration (13,16,27). Between February and June 2021, we reported waning of RSV neutralizing antibodies in infants and women of childbearing age in the Vancouver metropolitan area after the prolonged absence of viral exposure (16). The lack of worsening in RSV outcomes notwithstanding these observations may be reassuring. However, further monitoring is required in case maternal boost and placental antibody transfer were affected by the low RSV circulation with potential lag in the clinical implications for newborns yet to be fully realized. Our findings reinforce the importance of active surveillance and individual reportability of RSV cases to systematically assess resurgence and to guide recommendations for prompt initiation and who should immunoprophylaxis in children most susceptible to severe disease.

Our study has limitations mainly related to the surveillance and observational nature of our data. All surveillance data are subject to incomplete, missing, or inaccurate information and as for all observational designs, our analyses are subject to bias and artefact, including related to healthcare seeking behaviours among parents/children and testing practices/proclivities among clinicians including systematic changes during the pandemic as we have detailed here. Pre-pandemic, children with mild infections were often not tested for RSV contributing to underestimation of the number of RSV cases. In addition to the more liberal testing criteria for RSV and other respiratory virus for children being admitted to hospital during the 2021-22 season, there may have been changes in health seeking behaviours as well as clinician testing practices in the wake of the pandemic. The broadened testing criteria specifically for admitted patients may have resulted in increased RSV hospitalization detection during the pandemic period. Finally, differences between jurisdictions (e.g., population density, social behaviors, seasonal factors, etc.) warrants national data to get a more complete picture.

In conclusion, RSV circulation halted during the pandemic, but with the lifting of mitigation measures a subsequent resurgence among children <36 months of age was accompanied by shift toward older (24-35 month) cases in 2021-22, without increased severity. For the 2022-23 season, increased circulation, residual vulnerability in additional birth cohorts also spared RSV infection during the pandemic as well as a deteriorated access to primary care could have marked cumulative impact with with serious implications for healthcare systems and capacity, even if the same proportion are hospitalized. This study provides important clinical severity data, otherwise limited to date in Canada, and highlights the importance of ongoing systematic and strategic RSV surveillance, inclusive of severe outcome monitoring, to optimize preparation and prevention approaches.

## Data Availability

All data produced in the present work are contained in the manuscript.

## Acknowledgements

We thank Cheryl Christopherson and Grace Burns, from the BC RSV Immunoprophylaxis Program, and Alireza Saremi and Henry Cho, from Provincial Health Services Authority, for providing data, Dr. Christina Michalski for input in study design and editing of the manuscript.

## Tables and figures

**Supplemental figure 1:**
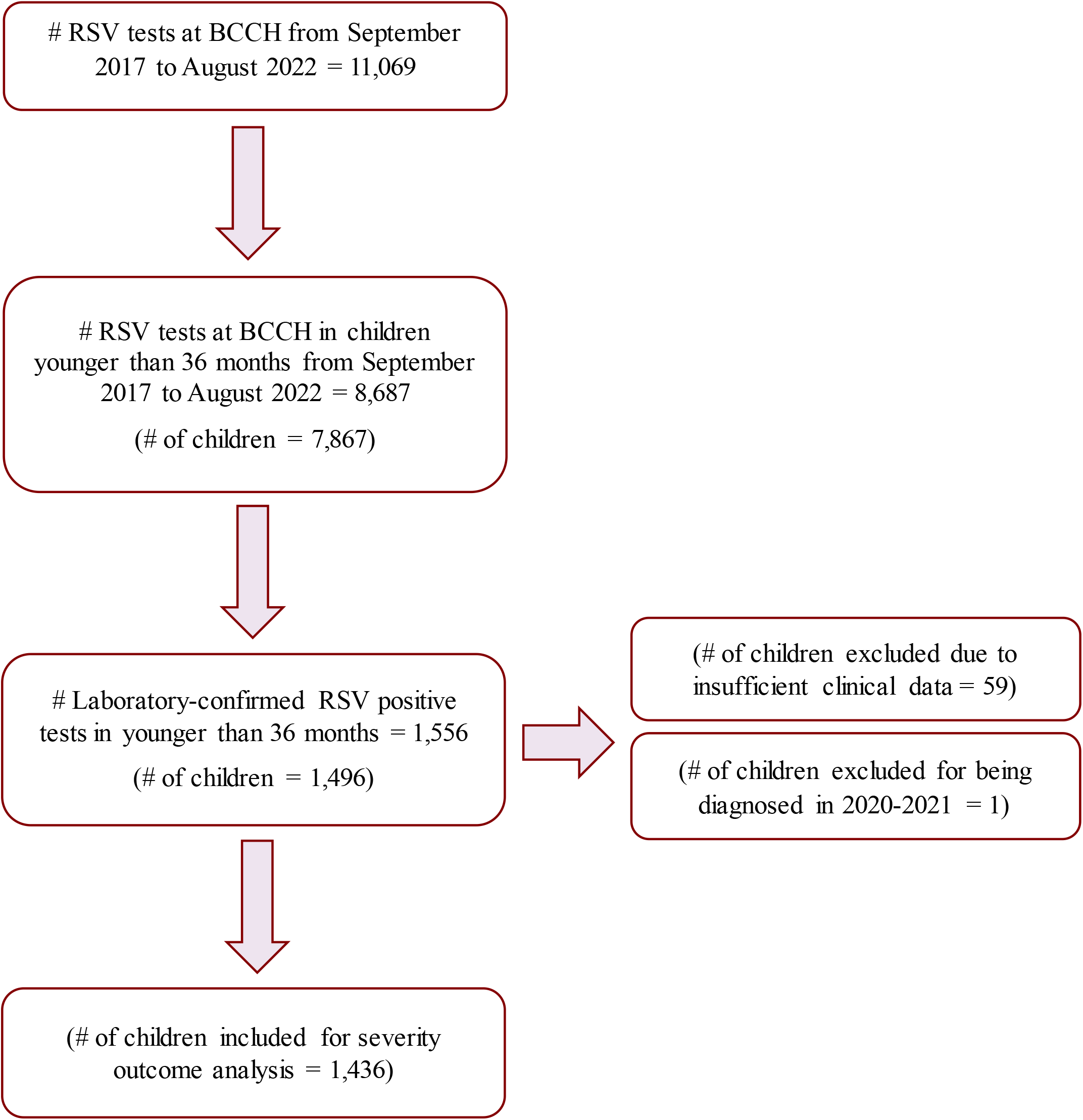
Flowchart of RSV cases included in study.

**Supplemental figure 2:**
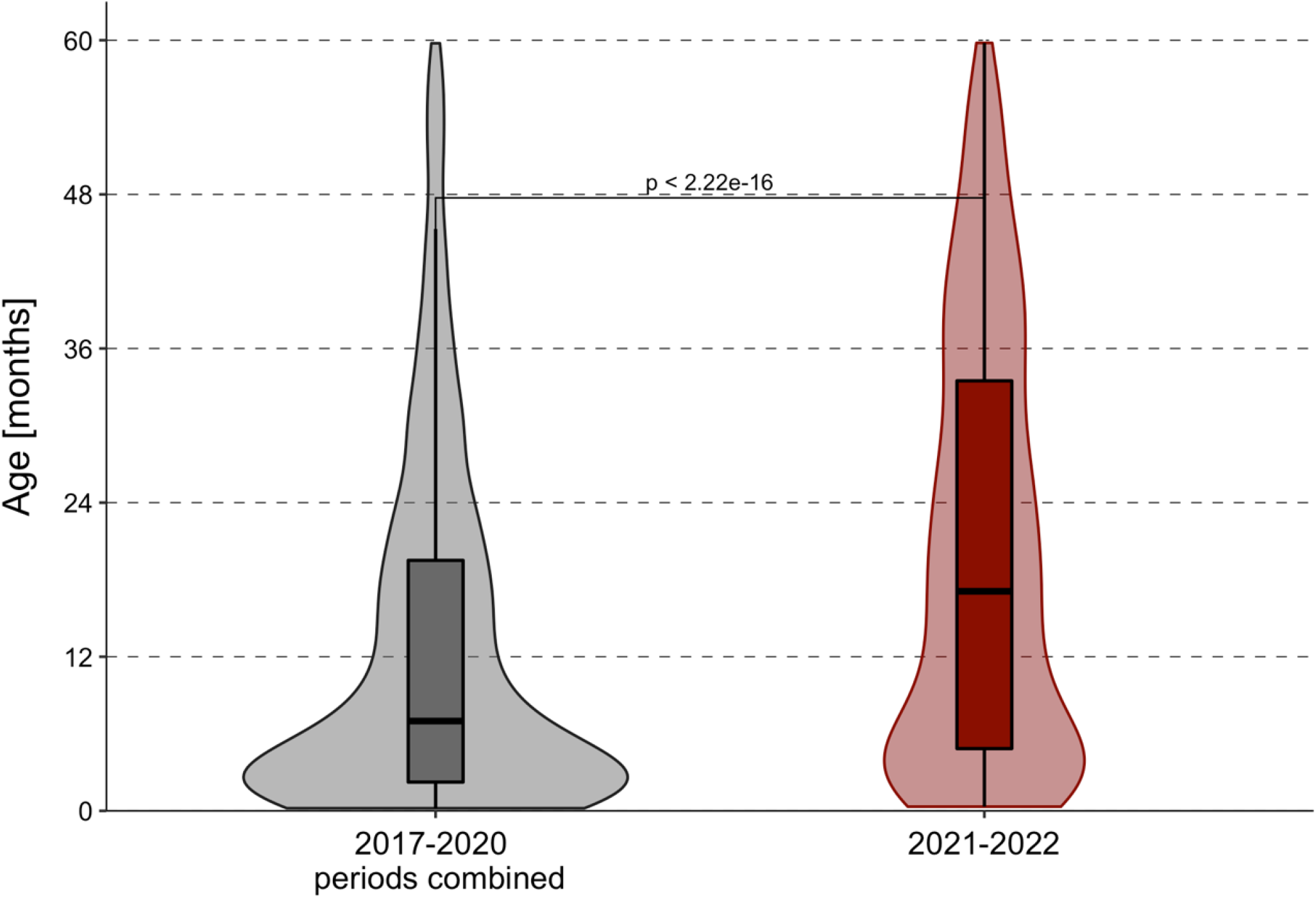
Age distribution of children diagnosed with RSV at BCCH. Combined data from September 1^st^ 2017 to August 31^st^ 2020 (grey area), and September 1^st^ 2021 to August 31^st^ 2022 (red area); statistical comparisons by Mann Whitney U test. Data includes RSV cases <18-year-old.

**Supplemental table 1:**
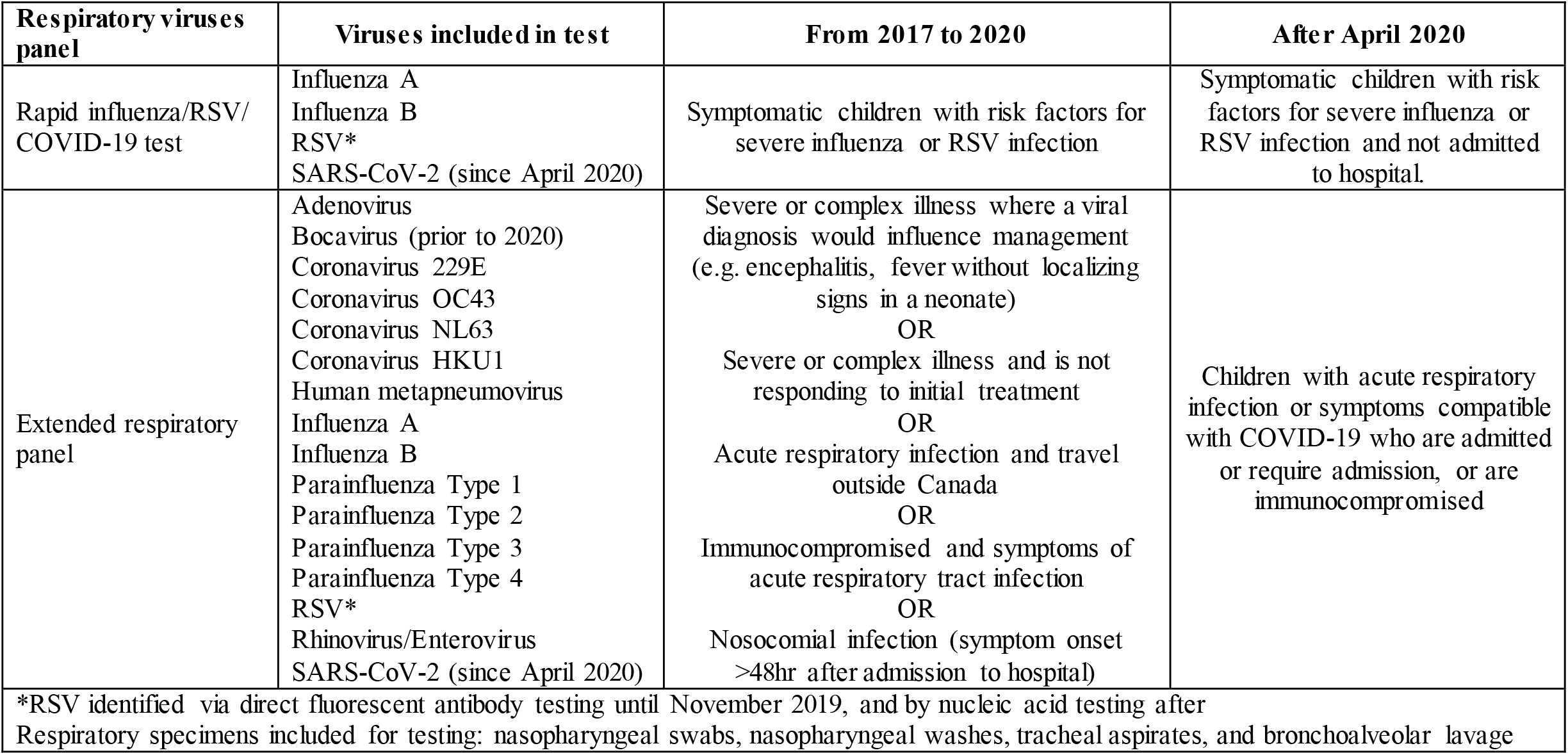
Indication for testing by respiratory virus panels at the BCCH Microbiology Laboratory.

| **Respiratory viruses panel** | **Viruses included in test** | **From 2017 to 2020** | **After April 2020** |
| --- | --- | --- | --- |
| Rapid influenza/RSV/COVID-19 test | Influenza A<br>Influenza B<br>RSV*<br>SARS-CoV-2 (since April 2020) | Symptomatic children with risk factors for severe influenza or RSV infection | Symptomatic children with risk factors for severe influenza or RSV infection and not admitted to hospital. |
| Extended respiratory panel | Adenovirus<br>Bocavirus (prior to 2020)<br>Coronavirus 229E<br>Coronavirus OC43<br>Coronavirus NL63<br>Coronavirus HKU1<br>Human metapneumovirus<br>Influenza A<br>Influenza B<br>Parainfluenza Type 1<br>Parainfluenza Type 2<br>Parainfluenza Type 3<br>Parainfluenza Type 4<br>RSV*<br>Rhinovirus/Enterovirus<br>SARS-CoV-2 (since April 2020) | Severe or complex illness where a viral diagnosis would influence management (e.g. encephalitis, fever without localizing signs in a neonate)<br>OR<br>Severe or complex illness and is not responding to initial treatment<br>OR<br>Acute respiratory infection and travel outside Canada<br>OR<br>Immunocompromised and symptoms of acute respiratory tract infection<br>OR<br>Nosocomial infection (symptom onset >48hr after admission to hospital) | Children with acute respiratory infection or symptoms compatible with COVID-19 who are admitted or require admission, or are immunocompromised |
| *RSV identified via direct fluorescent antibody testing until November 2019, and by nucleic acid testing after<br>Respiratory specimens included for testing: nasopharyngeal swabs, nasopharyngeal washes, tracheal aspirates, and bronchoalveolar lavage |  |  |  |

**Supplemental table 2:**
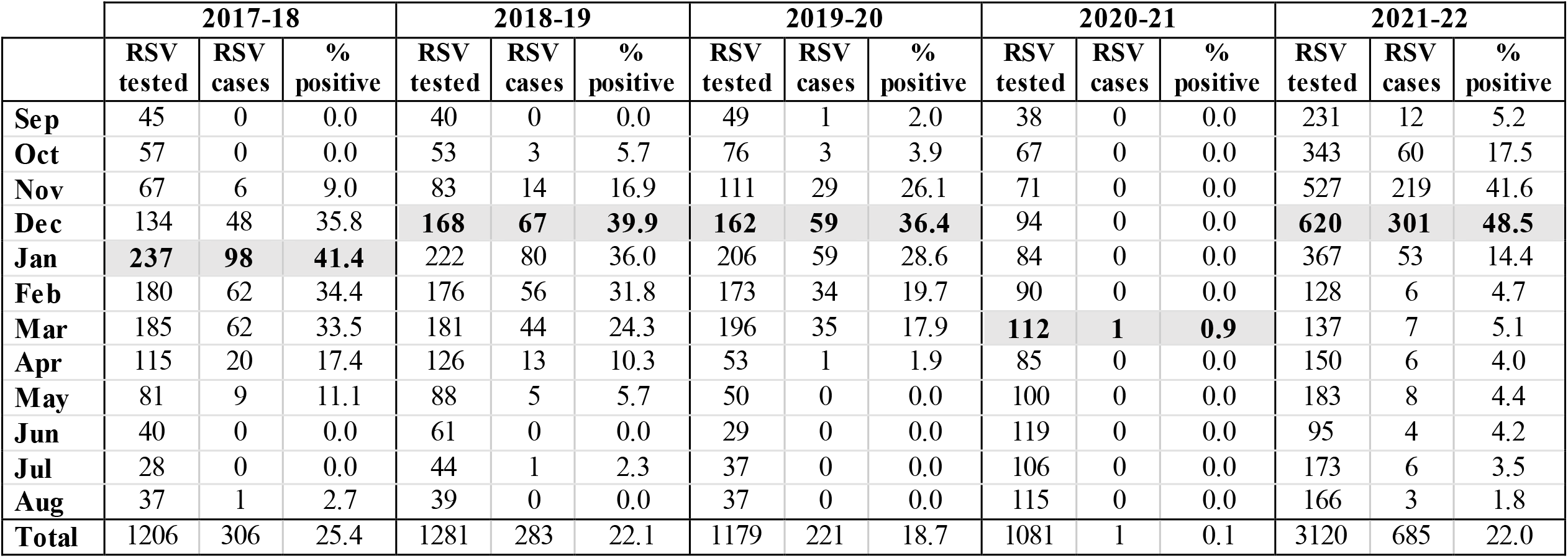
Monthly RSV tests, cases and testing positivity rate in children younger than 36 months at BCCH, by n period. Dates refer to time of collection of respiratory specimens. Peak monthly positivity rate for each period is highlighted in bold.

**Supplemental table 3:** Hospitalization and severity outcomes by age groups and annual periods, for children with RSV at BCCH.

|  | September 1<sup>st</sup> to August 31<sup>st</sup> period<br>(number of RSV cases; n = 1,436) |  |  |  |  |
| --- | --- | --- | --- | --- | --- |
| Age group | 2017-18<br>(n = 259) | 2018-19<br>(n = 273) | 2019-20<br>(n = 219) | 2017-20 periods<br>combined (n = 250)<sup>1</sup> | 2021-22<br>(n = 685) |
| Severity outcomes |  |  |  |  |  |
| **<6 months old, n (%)<sup>2</sup>** | 135 (52.1) | 133 (48.7) | 103 (47.0) | 124 (49.4) | 241 (35.2) |
| Hospitalized, n (% age group)<sup>3</sup> | 74 (54.8) | 57 (42.9) | 52 (50.5) | 61 (49.3) | 63 (26.0) |
| Length of in-hospital stay, median days (IQR) | 4 (2 - 6) | 4 (2 - 5) | 4 (2 - 7) | 4 (2 - 6) | 3 (2 - 6) |
| Received supplemental O<sub>2</sub>, n (% hospitalized) | 49 (66.2) | 39 (68.4) | 39 (75.0) | 43.25 (70.3) | 46 (73.0) |
| ICU admission, n (% hospitalized) | 18 (24.3) | 10 (17.5) | 13 (25.0) | 14.25 (23.2) | 16 (25.4) |
| **6 to <12 months old, n (%)<sup>2</sup>** | 41 (15.8) | 40 (14.7) | 33 (15.1) | 38 (15.2) | 106 (15.5) |
| Hospitalization, n (% age group) <sup>3</sup> | 25 (61.0) | 16 (40.0) | 20 (60.6) | 20 (53.5) | 13 (12.3) |
| Length of in-hospital stay, median days (IQR) | 3 (1 - 5) | 4 (3 - 5) | 3 (2 - 4) | 3 (2 - 5) | 4 (3 - 6) |
| Received supplemental O<sub>2</sub>, n (% hospitalized) | 18 (72.0) | 11 (68.8) | 13 (65.0) | 13 (70.3) | 10 (76.9) |
| ICU admission, n (% hospitalized) | 4 (16.0) | 4 (25.0) | 1 (5.0) | 3 (16.2) | 3 (23.1) |
| **12 to <24 months old, n (%)<sup>2</sup>** | 53 (20.5) | 69 (25.3) | 57 (26.0) | 60 (23.8) | 188 (27.4) |
| Hospitalization, n (% of age group) <sup>3</sup> | 32 (60.4) | 48 (69.6) | 32 (56.1) | 37 (62.6) | 23 (12.2) |
| Length of in-hospital stay, median days (IQR) | 3 (2 - 6) | 3(2 - 5) | 3 (2 - 4) | 3 (2 - 5) | 2 (2 - 4) |
| Received supplemental O<sub>2</sub>, n (% hospitalized) | 22 (68.8) | 31 (64.6) | 18 (56.2) | 22.25 (65.9) | 18 (78.3) |
| Intensive care admission, n (% hospitalized) | 7 (21.9) | 7 (14.6) | 7 (21.9) | 6.25 (18.5) | 4 (17.4) |
| **24 to <36 months old, n (%)<sup>2</sup>** | 30 (11.6) | 31 (11.4) | 26 (11.9) | 29 (11.6) | 150 (21.9) |
| Hospitalization, n (% age group) <sup>3</sup> | 22 (73.3) | 17 (54.8) | 11 (42.3) | 17 (57.5) | 24 (16.0) |
| Length of in-hospital stay, median days (IQR) | 4 (3 - 9) | 4 (3 - 6) | 3 (2 - 6) | 4 (3 - 7) | 3 (2 - 4) |
| Received supplemental O<sub>2</sub>, n (% hospitalized) | 13 (59.1) | 12 (70.6) | 5 (45.5) | 11.75 (63.5) | 17 (70.8) |
| ICU admission, n (% hospitalized) | 8 (36.4) | 4 (23.5) | 1 (9.1) | 4.25 (23.0) | 4 (16.7) |
| Total hospitalized (SD) | 153 | 138 | 115 | 135 (15.6) | 123 |

<sup>1</sup> N (%) corresponds to average number per annual period, combining data from entire 3-annual period except for length of in-hospital stay where median and IQR were calculated from all values from the 2017-20 period;
<sup>2</sup> Proportion of that age subgroup out of all cases in that period (e.g., in 2017-18 for <6 months: 135/259);
<sup>3</sup> Proportion of hospitalized cases over total cases in that age group, per period (e.g., in 2017-18 for <6 months: 74/135).

**Supplemental table 4:** Need for respiratory therapy in children under 36 months of age with RSV at BCCH, by one-year period.

|  | September 1<sup>st</sup> to August 31<sup>st</sup> period |  |  |  |
| --- | --- | --- | --- | --- |
|  | 2017-18<br>(n = 259) | 2018-19<br>(n = 273) | 2019-20<br>(n = 219) | 2021-22<br>(n = 685) |
| Hospitalized, n (%) | 153 (59.1) | 138 (50.5) | 115 (52.5) | 123 (18.0) |
| Received supplemental O<sub>2</sub> or respiratory support, n (% hospitalized) | 102 (66.6) | 93 (67.4) | 75 (65.2) | 91 (74.0) |
| Blow by, n (% hospitalized) | 10 (6.5) | 4 (2.9) | 1 (0.9) | 10 (8.1) |
| Nasal prongs / low flow O<sub>2</sub>, n (% hospitalized) | 19 (12.4) | 19 (13.8) | 14 (12.2) | 20 (16.3) |
| High flow, n (% hospitalized) | 32 (20.9) | 35 (25.4) | 38 (33.0) | 38 (30.9) |
| BiPAP, n (% hospitalized) | 22 (14.4) | 20 (14.5) | 15 (13.0) | 21 (17.1) |
| CPAP, n (% hospitalized) | 1 (0.7) | 0 (0) | 1 (0.9) | 2 (1.6) |
| Not specified, n (% hospitalized) | 18 (11.8) | 15 (10.9) | 6 (5.2) | 0 (0) |

**Supplemental table 5:** Multivariate analysis of RSV-re late d hospitalization for co-morbidity groups versus no co-morbidities. Adjusted logistic regression model for RSV-related hospitalization with types of morbidity as predictors (used as binomial variables; i.e., presence of that comorbidity or no presence of that comorbidity).

|  | **2017-20 periods**<br>**combined (N=604)** | **2021-22 (N=463)** |
| --- | --- | --- |
| **Type of co-morbidity** | **OR (95%CI)** | **OR (95% CI)** |
| Respiratory | 6.54 (3.09 - 16.1) | 8.05 (2.84 - 24.54) |
| Cardiac | 2.03 (0.83 - 5.44) | 1.61 (0.36 - 5.38) |
| Neurologic | 1.81 (0.80 - 4.34) | 3.58 (0.90 -12.78) |
| Genetic / metabolic | 4.07 (1.28 – 18.0) | 2.68 (0.70 - 8.71) |

